# Association of Skeletal Muscle Function Deficit (SMFD) with Performance, Disability, Frailty Phenotype, Hospitalization, and Falls: Results from the *InCHIANTI* Study

**DOI:** 10.1101/2025.10.06.25337404

**Authors:** Angelo Di Iorio, Matteo Candeloro, Roberto Paganelli, Raffaello Pellegrino, Giada Mariano, Stefania Bandinelli, Toshiko Tanaka, Luigi Ferrucci

## Abstract

**Background:** Age-related muscle dysfunction is a major contributor to disability, frailty, and poor clinical outcomes in older adults. Muscle mass and strength provide limited insight into the multifactorial nature of muscular decline. Skeletal Muscle Function Deficit (SMFD) framework integrates multiple domains muscle mass, quality, strength, and power to capture a broader spectrum of age-related muscle dysfunction.

**Objective:** To develop and validate a composite SMFD score and evaluate its association with key geriatric outcomes in older adults.

**Methods:** This study used data from the InCHIANTI longitudinal cohort (1998–2018), including 1,035 participants and 3,196 total assessments. The SMFD score (range 0–20) was computed by assigning quintile-based values of muscle area, density, strength, and lower limb power. Associations with disability in basic and instrumental activities of daily living (BADL/IADL), frailty phenotype, poor physical performance (SPPB <7), hospitalization, falls number, and major chronic diseases were analyzed using mixed-effects models, adjusting for age, sex, fat area, and multimorbidity.

**Results:** The SMFD score declined significantly over time and was independently associated with lower risk of BADL (OR 0.57), IADL (OR 0.70), frailty (OR 0.72), poor performance (OR 0.68), hospitalization (OR 0.96), and falls number (OR 0.96). Higher SMFD scores were also inversely associated with the prevalence and incidence of Parkinson’s disease, stroke, and hip osteoarthritis.

**Conclusions:** The SMFD score is a valid, multidimensional measure that predicts adverse outcomes in older adults more effectively than traditional sarcopenia, dynapenia, and powerpenia. It holds promise for use in clinical assessment, risk stratification, and targeted interventions.

## 1.0 Introduction

The aging process is associated with a decline in musculoskeletal function, which significantly impacts the quality of life, functional independence, and survival of older adults (Ferrucci et al., 2000). Among these conditions, Sarcopenia has gained increasing acknowledgement. Originally defined by Rosenberg as the age-related loss of skeletal muscle mass, sarcopenia was conceptually and operationally revised (Costanzo et al., 2020; Rosenberg and Roubenoff, 1995). Early definitions highlighted a quantitative reductions in lean body mass, whereas was consistently demonstrated that muscle mass alone does not fully explain the observed impairments in strength, mobility, or risk of adverse outcomes in the elderly (Correa-De-Araujo and Hadley, 2014). The introduction of operational criteria, by the European Working Group on Sarcopenia in Older People (EWGSOP) (Cruz-Jentoft et al., 2019) and the Foundation for the National Institutes of Health (FNIH) Sarcopenia Project (Studenski et al., 2014), integrate in the definition low muscle mass, low strength and/or poor physical performance. Despite this improvement in the operational definition, important challenges remain in capturing the multifactorial nature of muscle dysfunction in aging. For instance, fatigue and functional limitations may be reported even in subjects with preserved muscle mass, suggesting a disconnection between quantity and quality in muscle aging (Cooper et al., 2015).

Dynapenia was proposed to disentangle specific deficits in muscle function, and describes the age-related loss of muscle strength, irrespective of muscle mass (Clark and Manini, 2008). Both Sarcopenia and dynapenia independently predict disability (Clark and Manini, 2010), falls, hospitalization (Visser and Schaap, 2011), and mortality (Clark and Manini, 2012). Notably, muscle power reduction (powerpenia, defined as the reduction in the product of the force contraction and velocity of movement) may occur earlier in the life span, and could progress more rapidly than the reduction in mass (Freitas et al., 2024), and is influenced by several aspects similar to sarcopenia and dynapenia, as neuromuscular, metabolic, and structural factors (Pellegrino et al., 2025).

Moreover, muscle quantity measures alone do not capture the interplay of age-related changes that together influence mobility and metabolic health.

The term Skeletal Muscle Function Deficit (SMFD) has been proposed (Correa-De-Araujo and Hadley, 2014) to bridge the gap between those limitations and to provide a more encompassing clinical construct. As matter of fact, SMFD captures the spectrum of age-related muscle dysfunctions, encompassing sarcopenia, dynapenia, powerpenia, inflammation and myosteatosis (fat muscle infiltration) (Correa-De-Araujo and Hadley, 2014). SMFD reflects a shift from an exclusively descriptive perspective to a more comprehensive vision of aging health.

This broader conceptualization allows clinicians and researchers to move beyond the limitations of single-criterion definitions and recognize the heterogeneity of muscle dysfunction in older adults. It aligns with contemporary models of chronic disease, such as heart failure or COPD, where diagnosis encompasses functional impairments, symptom burden, and systemic involvement.

Despite this intriguing conceptualization, are missing evidence that SMFD may offer superior predictive validity for relevant geriatric outcomes, including disability in activities of daily living (ADL), mobility decline, hospitalization, and falls, compared to traditional metrics based solely on muscle mass and strength.

Our hypothesis is that: a summary measure of the different components of the SFMD (power, strength, density and area of the muscle), by capturing the multidimensional nature of muscle dysfunction, may serve as a more accurate and operationally useful tool for clinical assessment and research.

Therefore the study aims to propose a summary score of the SFMD, and to validate in predicting adverse health outcomes in older adults such as decline in activities of daily living, physical performance, frailty phenotype, number of falls, risk of hospitalization, and major chronic diseases.

## 2.0 Methods

The InCHIANTI study protocol has been extensively detailed elsewhere with regard to its operational procedures (Ferrucci et al., 2000). The study design and research implementation were developed by the Laboratory of Clinical Epidemiology at the National Institute of Research and Care of Aging (INRCA, Florence).

The study protocol was approved by the Italian National Institute of Research and in the United States the protocol was given an exemption status by the National Institutes of Health Intramural Research Program Institutional Review Board (Exemption #11976) and informed consent was obtained from participants at each visit.

The study started between 1998 and 2000 and included triennial follow-ups, concluding in 2018. It was conducted at two distinct Italian sites: a suburban area near Florence and a rural area in the Chianti region. A total of 1,453 participants were enrolled. For the present analyses, 1,035 baseline participants were included, yielding a total of 3,196 longitudinal observations across the follow-up waves. For this study data from the baseline were utilized, through 3 follow-ups, as all variables of interest were available.

### 2.1 Muscle assessment

#### 2.1.1 Tibial peripheric quantitative computed tomography

The peripheral quantitative computed tomography (pQCT) was performed by the XCT 2000 device (Stratec Medizintechnik, Pforzheim, Germany) (Russo et al., 2003). The images obtained from the pQCT were analyzed using the BonAlyse software (BonAlyse Oy, Jyvaskyla, Finland) (Capozza et al., 2010). The following bone parameters were derived from the pQCT images measured at 38% tibia length (Russo et al., 2006): calf muscle cross-sectional area (cm^2^) was evaluated at 38% of the tibia length from the distal tip of the tibia; whereas fat cross-sectional area (cm^2^), muscle density (mg/cm^3^) at a transverse scan performed at 66% of the tibia length.

#### 2.1.2 Muscle strength

Grip strength was measured with a handheld dynamometer (hydraulic hand BASELINE; Smith and Nephew). Participants were asked to perform the task twice with each hand, and the maximum strength attained during the four trials was used for the present analysis.

#### 2.1.3 Lower limb muscle power evaluation

Maximal voluntary leg extension power was evaluated with the Nottingham power rig (Medical Engineering Unit, University of Nottingham Medical School, Nottingham, UK) (Bassey and Short, 1990) according to Leg Extension Power Rig user manual.

The participants were seated in an upright position, arms folded across the chest, and knees flexed with one foot placed on the floor and the other foot placed on the dynamometer pedal. The sitting position was determined so that the knee reached 15° of flexion (0° is full extension) at the end of the footplate movement (Bassey and Short, 1990; Lauretani et al., 2003). Participants familiarized with the procedure in two warm-up trials and then instructed to push the pedal forward as hard and fast as possible. Measurements were repeated for each limb until maximal power could not be increased further. The highest lower limb muscle power was selected for further analysis (Lauretani et al., 2003).

#### 2.1.4 Skeletal Muscle Function Deficit

Based on the SMFD definition proposed by Araujo, we categorized lower limb muscle mass, muscle quality, strength, and explosive power into quintiles of distribution, assigning a score from one (lowest quintile) to five (highest quintile). Participants who, during any phase of the study, were unable to perform or could not be assessed for one of the parameters were assigned a score of zero. Accordingly, the total SMFD score ranged from 0 to 20.

### 2.2 Physical Performance and strength

The Short Physical Performance Battery (SPPB) is a tool used to assess lower extremity performance, based on the Established Populations for the Epidemiologic Studies of the Elderly (EPESE) (Guralnik et al., 1995). SPPB consists of three tests: walking speed, ability to stand from a chair, and ability to maintain balance in progressively more challenging positions. Each test is scored on a five-level scale, with 0 indicating inability to perform the test and 4 indicating the highest level of performance. The scores from the three tests are then added together to create a summary physical performance measure, ranging from 0 (worst) to 12 (best).

### 2.3 Measurement of functional ability

At baseline and at every follow-up visit, basic activities of daily living (BADL) and instrumental (IADL) were assessed and a summary score was calculated as the number of impaired BADL and IADL. Moreover two or more reported disabilities were considered as the cut-off to define IADL and/or BADL as a disabled subject.

### 2.4 Frailty phenotype

Frailty phenotype was defined according to Fried definition, and assessed in every time of the study, as the presence of at least 3 of the following domains (Fried et al., 2001): 1) unintentional weight loss, a reduction in weight >4.5 kilograms in the past 12 months; 2) Exhaustion, defined as a feeling of needing an effort to do everything for more than 2 times in the week before interview; 3) Reduced physical activity, defined as having performed less than 2-4 hours of light exercise per week; 4) walking speed, defined as time needed to walk at usual pace for 4 m in the upper quintile; 5) Grip strength in the gender-specific bottom quintile.

### 2.5 Fall assessment

Both at baseline and in subsequent follow-ups, falls were assessed in quantitative and qualitative terms, and the information collected was verified with the help of caregivers and general practitioners.

### 2.6 Body composition

Body Mass Index (BMI, kg/m2) was calculated using objectively measured height and weight. Weight was measured to the nearest 0.1 kg using a high-precision mechanical scale and standing height to the nearest 0.1 cm with a wall measure with participants wearing light indoor clothes and no shoes (Ferrucci et al., 2000).

### 2.7 Diseases and multimorbidity

Based on self-report, the diagnosis of major medical conditions was ascertained according to preestablished criteria that combine reported doctor diagnosis, eventually supported by medical records, physical examination, blood tests and drugs prescription (Simonsick et al., 1997).

Multimorbidity score was calculated summing the number of diseases reported at baseline and all follow-up visits (angina, cancer, hepatic diseases, acute myocardial infarction, congestive heart failure, stroke, Parkinson disease, peripheral artery disease, diabetes, COPD, asthma, and arthrosis).

### 2.7 Statistical analysis

Baseline characteristics were compared between times of the study for all variables of interest using analysis of variance for continuous variables and for trend χ^2^ test analyses for dichotomous or categorical variables. Moreover, were also reported differences in demographic and clinical features of the InCHIANTI study population according to population quintile of distribution in: lower limb muscle mass, muscle quality, strength, and explosive power, using analysis of variance for continuous variables and for trend χ^2^ test analyses.

Hierarchical Generalized Linear Models (HGLM, Proc GLIMMIX) models were used to specify a repeated measures model with a dichotomous outcome variable (binary logistic regression) and multiple independent factors (Ene et al., 2015). The different models encompass as outcome variables: disability in basic and instrumental ADL (score 2 or more for both), low physical performance (SPPB <7), Frailty phenotype (Fried score >2), hospitalization. Conversely to assess predictors of the number of falls, HGLM was also used but with Poisson distribution.

As independent predictors were considered, chronological age, sex, multimorbidity score (as the sum of every disease reported in the different study assessment), fat area measured at the 66% of tibia length, and SMFD-score.

To test whether SFMD-score could predict also incident and prevalent diseases compared to those subjects that do not report the specific disease, GEE repeated measures model, with ordinal option, was also used, the model consider as confounders the same variables reported in previous analysis (chronological age, sex, multimorbidity, and fat area measured at the 66% of tibia length).

SAS version 9.4 for Windows (SAS Institute, Inc., Cary, NC) was used for all data processing and statistical analyses. We set the level of statistical significance at p < 0.05 (2-sided).

## 3.0 Results

Over the course of the InCHIANTI longitudinal study, involving an initial cohort of 1035 older participants, with a total of 3196 assessments, a progressive and significant deterioration of muscle function and overall health status was observed. Across the follow-up waves, the SFMD-score demonstrated a marked and continuous decline, decreasing from an average value of 9.60±4.20 at baseline to 7.07±4.00 at the third follow-up visit (p < 0.001) (Table 1). This steady decline reflects the compounded deterioration of both functional and morphological aspects of skeletal muscle integrity. In parallel with this muscular decline, participants’ mean chronological age increased from 75.5±7.39 to 80.4±5.99 years, accompanied by a notable rise in multimorbidity, with the average number of chronic conditions increasing from 2.15±1.53 to 3.42±1.72 (for trend p < 0.001).

**Table 1:** Demographic and clinical features of the InCHIANTI study population according to the times of the study.

|  | Baseline | FU-1 | Fu-2 | Fu-3 | p-value |
| --- | --- | --- | --- | --- | --- |
|  | 1035 | 835 | 739 | 587 |  |
| Chronological age (yy) | 75.48±7.39 | 77.18±7.01 | 78.97±6.60 | 80.41±5.99 | <0.001 |
| Age at baseline (yy) | 74.97±7.39 | 73.62±7.01 | 72.36±6.60 | 70.74±5.98 | <0.001 |
| Male sex n (%) | 458 (44.25) | 365 (43.71) | 311 (42.08) | 251 (42.76) | 0.19 |
| SFMD score (0-20) | 9.60±4.20 | 9.55±4.73 | 8.70±4.61 | 7.07±4.00 | <0.001 |
| BADL score (0-6) | 0.26±1.02 | 0.41±1.29 | 0.47±1.40 | 0.49±1.45 | <0.001 |
| BADL disability n (%) | 58 (5.60) | 74 (8.86) | 77 (10.42) | 61 (10.39) | <0.001 |
| IADL score (0-8) | 0.96±2.15 | 1.14±2.37 | 1.75±2.67 | 1.42±2.51 | <0.001 |
| IADL disability n (%) | 140 (13.53) | 135 (16.17) | 187 (25.30) | 120 (20.44) | <0.001 |
| SPPB score (0-12) | 9.72±3.29 | 9.10±3.59 | 8.80±3.76 | 9.35±3.43 | <0.001 |
| SPPB score < 7 n (%) | 216 (20.87) | 207 (24.79) | 206 (27.88) | 159 (27.09) | <0.001 |
| Fried Frailty Phenotype | 85 (8.21) | 85 (10.18) | 91 (12.31) | 82 (13.97) | 0.001 |
| Multimorbidity score | 2.15±1.53 | 2.68±1.71 | 3.03±1.77 | 3.42±1.72 | <0.001 |
| Falls mean number | 0.41±0.99 | 0.42±0.95 | 0.42±1.12 | 0.40±1.15 | 0.90 |
| Faller n (%) | 236 (22.80) | 198 (23.71) | 170 (23.00) | 118 (20.10) | 0.26 |
| Muscle area (cm<sup>2</sup>) | 62.29±12.65 | 64.02±12.67 | 60.56±12.33 | 58.10±12.37 | <0.001 |
| Muscle density (mg/cm<sup>3</sup>) | 70.56±3.74 | 69.86±3.32 | 70.29±3.49 | 69.63±3.57 | <0.001 |
| Muscle Strength (Kg) | 28.57±11.95 | 30.74±11.58 | 29.05±11.82 | 25.25±9.81 | <0.001 |
| Muscle Power (W) | 101.51±62.41 | 122.86±62.71 | 99.19±56.23 | 86.94±46.24 | <0.001 |
| Fat area (cm<sup>2</sup>) | 19.14±11.70 | 19.31±11.79 | 20.19±11.28 | 20.71±11.61 | 0.008 |
| Body Mass Index | 27.46±4.11 | 26.47±3.98 | 27.18±4.14 | 27.15±4.17 | 0.87 |

Functional disability also worsened progressively: basic activities of daily living (BADL) disability prevalence rose from 5.6% to 10.4%, while instrumental activities of daily living (IADL) disability increased from 13.5% to 20.4% (both for trend-p < 0.001). Physical performance, as measured by the Short Physical Performance Battery (SPPB), showed a biphasic trajectory: an initial decline from 9.72±3.29 to 8.80±3.76, followed by a partial recovery to 9.35±3.43 at the final follow-up (for trend p < 0.001), reflecting individual variability in functional trajectories. Furthermore, frailty, as assessed by Fried’s phenotype, increased from 8.2% to 14% over the study period (p = 0.001), underscoring the progressive vulnerability of the aging cohort.

Detailed analysis of muscular components revealed a consistent deterioration across multiple domains: Muscle power, Handgrip strength, Muscle cross-sectional area, and Muscle density decreased statistically significant across time of the study. In contrast, BMI and the fat area remained relatively stable throughout the observation period. These findings suggest that the observed muscle decline reflects a qualitative deterioration in muscle tissue, rather than gross changes in body weight or fat distribution.

In Figure 1 and Figure 2, the transitions analysis between quintiles of distribution in leg muscle power, muscle area, muscle density, muscle strength and SMFD-score, across time of the study is reported. Synthetically, as could be expected, subjects transitioned from higher to lower performance quintiles in all muscle parameters, across time of the study, and is evident the accumulation in the cluster “unable to perform” specific test before dying. Lastly only a very small subset of subjects maintained stable high-performance levels.

**Figure 1:**
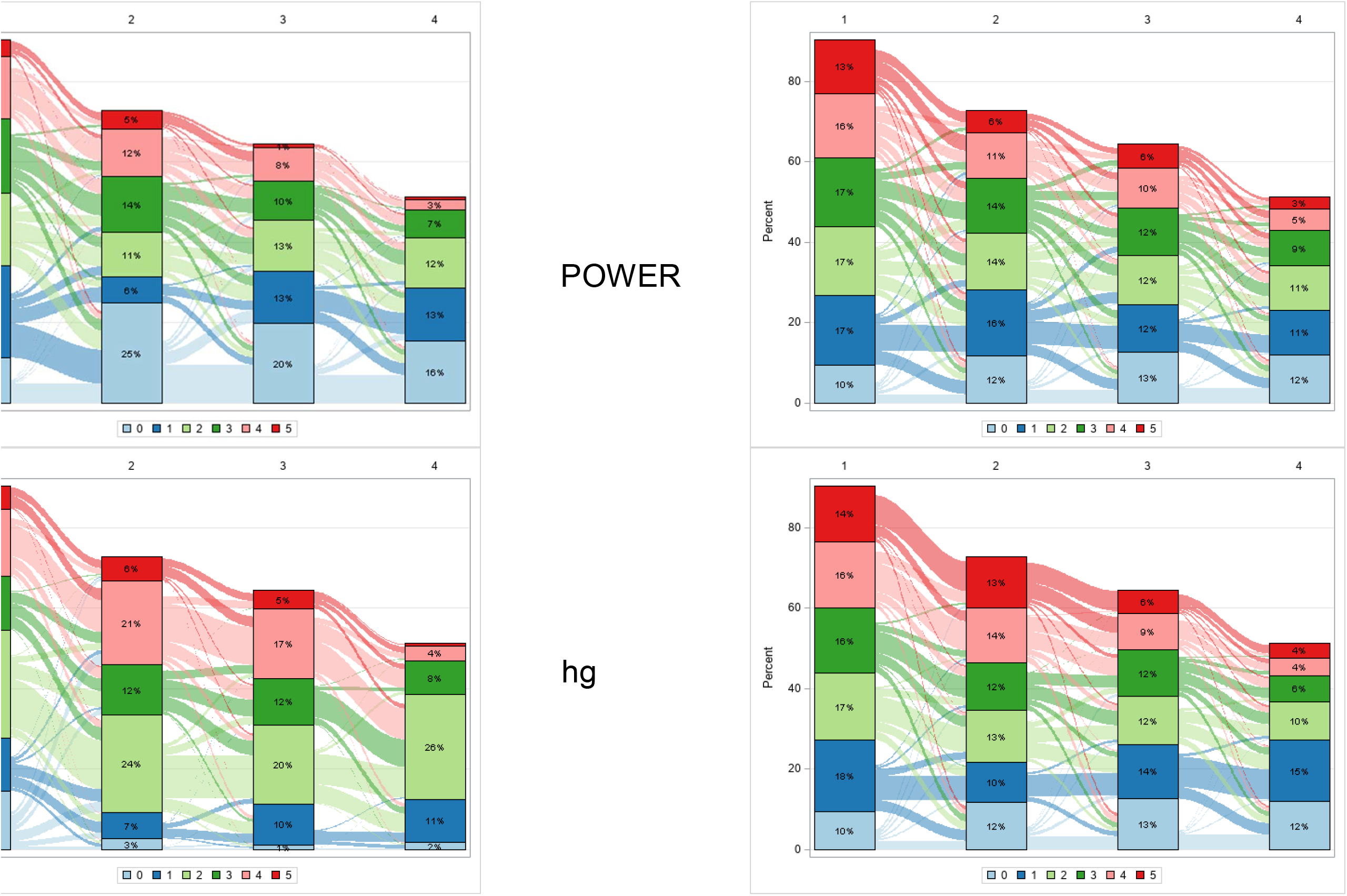
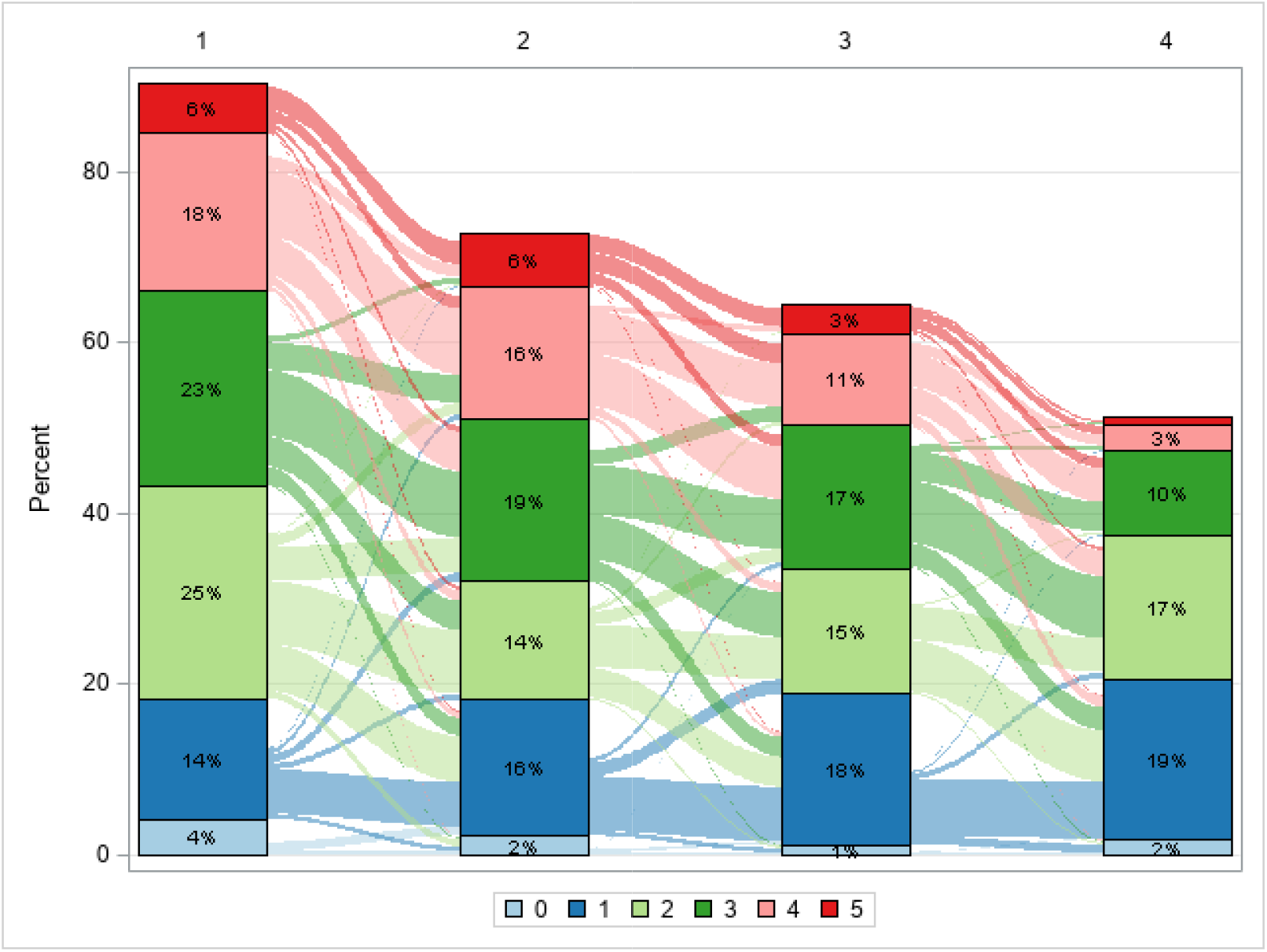
Sankey graphs reporting transition between quintiles of distribution in muscle power of lower limb (Figure 1 A), muscle grip strength (Figure 1 B), muscle density (Figure 1 C), and muscle mass (Figure 1 D). Legend: Group 0, subjects unable to do or to perform; Group 1-5, quintiles of distribution.

In Supplementary Table 1-4, are reported: demographic and clinical features of the study population according to quintile distribution of muscle markers. Higher quintiles of distribution for every muscle-markers were statistically significant associated with younger age, higher SFMD-score, lower rates of multimorbidity, lower rate of disability in both BADL and IADL, and higher SPPB-score.

Logistic Mixed Models for repeated measures were used, to validate the predictive role of the SMFD-score, stratifying also for sex (Table 2 and Table 3). Higher SFMD-scores predicted lower odds of adverse clinical outcomes independently from age, sex and multimorbidity, in BADL (OR = 0.57; 95% CI: 0.46–0.69), IADL (OR = 0.70; 95% CI: 0.66–0.75), poor physical performance(SPPB < 7) (OR = 0.68; 95% CI: 0.64–0.73), Fried’s frailty phenotype (OR = 0.72; 95% CI: 0.68–0.76), hospitalization (OR = 0.96; 95% CI: 0.93–0.99), and falls’ number (OR = 0.96; 95% CI:0.92–0.99).

**Table 2:** GLIMMIX with a binomial distribution option, factors predicting Basic and InstrumentalADL disability, and low performance at the SPPB over the course of the study, and according to sex.

|  |  |  |  |
| --- | --- | --- | --- |
| ***Basic ADL*** | All | Male | Female |
|  | OR (95%CI) | OR (95%CI) | OR (95%CI) |
| Chronological age | 1.01 (0.93-1.09) | 0.95 (0.85-1.07) | 1.07 (0.97-1.18) |
| Sex (female vs male) | 0.92 (0.41-2.06) |  |  |
| Multimorbidity | **1.36 (1.04-1.79)** | **1.72 (1.21-2.44)** | 0.97 (0.74-1.28) |
| Fat area 66% | 1.00 (0.99-1.01) | 1.00 (0.99-1.01) | 1.00 (0.99-1.01) |
| SFMD score | **0.57 (0.46-0.69)** | **0.66 (0.53-0.82)** | **0.49 (0.37-0.64)** |
| ***Instrumental ADL*** | All | Male | Female |
|  | OR (95%CI) | OR (95%CI) | OR (95%CI) |
| Chronological age | **1.13 (1.08-1.17)** | **1.11 (1.04-1.17)** | **1.15 (1.11-1.20)** |
| Sex (female vs male) | **0.52 (0.34-0.78)** |  |  |
| Multimorbidity | **1.23 (1.12-1.36)** | 1.19 (0.99-1.41) | **1.27 (1.13-1.43)** |
| Fat area 66% | 1.00 (0.99-1.01) | 1.00 (0.99-1.01) | 1.00 (0.99-1.01) |
| SFMD score | **0.70 (0.66-0.75)** | **0.70 (0.62-0.78)** | **0.70 (0.65-0.75)** |
| ***Low performance*** | All | Male | Female |
|  | OR (95%CI) | OR (95%CI) | OR (95%CI) |
| Chronological age | **1.08 (1.06-1.10)** | **1.06 (1.02-1.10)** | **1.09 (1.05-1.14)** |
| Sex (female vs male) | **0.51 (0.36-0.73)** |  |  |
| Multimorbidity | **1.19 (1.10-1.28)** | **1.22 (1.06-1.40)** | **1.16 (1.05-1.28)** |
| Fat area 66% | 1.00 (0.99-1.01) | 1.00 (0.99-1.01) | 1.00 (0.99-1.01) |
| SFMD score | **0.68 (0.64-0.73)** | **0.64 (0.57-0.72)** | **0.70 (0.66-0.75)** |
| ***Frailty Phenotype*** | All | Male | Female |
|  | OR (95%CI) | OR (95%CI) | OR (95%CI) |
| Chronological age | **1.05 (1.02-1.08)** | 1.05 (0.99-1.11) | **1.05 (1.01-1.09)** |
| Sex (female vs male) | 0.76 (0.50-1.14) |  |  |
| Multimorbidity | **1.30 (1.18-1.43)** | **1.27 (1.11-1.46)** | **1.31 (1.16-1.47)** |
| Fat area 66% | 1.00 (0.99-1.01) | 1.00 (0.99-1.01) | 1.00 (0.99-1.01) |
| SFMD score | **0.72 (0.68-0.76)** | **0.67 (0.60-0.75)** | **0.74 (0.68-0.80)** |

**Table 3:** GLIMMIX with a binomial distribution option for factors predicting hospitalization, and a Poisson distribution option for number of falls over the course of the study, and according to sex.

| *Hospitalization* | All | Male | Female |
| --- | --- | --- | --- |
|  | OR (95%CI) | OR (95%CI) | OR (95%CI) |
| Chronological age | 1.20 (0.98-1.46) | **1.39 (1.05-1.84)** | 1.08 (0.83-1.42) |
| Sex (female vs male) | **1.77 (1.37-2.28)** |  |  |
| Multimorbidity | **1.19 (1.12-1.27)** | **1.16 (1.06-1.26)** | **1.23 (1.13-1.35)** |
| Fat area 66% | 0.99 (0.98-1.01) | **0.96 (0.94-0.99)** | 1.00 (0.99-1.02) |
| SFMD score | **0.96 (0.93-0.99)** | 0.99 (0.94-1.04) | **0.95 (0.90-0.99)** |
| *Falls Number* | All | Male | Female |
|  | OR (95%CI) | OR (95%CI) | OR (95%CI) |
| Chronological age | 1.12 (0.92-1.38) | 0.90 (0.61-1.34) | 1.21 (0.96-1.53) |
| Sex (female vs male) | **0.62 (0.47-0.81)** |  |  |
| Multimorbidity | **1.12 (1.06-1.18)** | 1.09 (0.99-1.19) | **1.14 (1.06-1.21)** |
| Fat area 66% | 1.00 (0.99-1.02) | 1.01 (0.99-1.05) | 1.01 (0.99-1.02) |
| SFMD score | **0.96 (0.92-0.99)** | **0.90 (0.83-0.98)** | 0.98 (0.94-1.01) |

The stratified analysis did not reveal any differences between the two sexes in the predictive models, except for the risk of falls and hospitalization. As a matter of fact, in male SMFD-score did not statistically significant predict hospitalization, and in female sex SMFD-score did not predict falls number.

Finally, GEE ordinal repeated measures, with a multinomial distribution were used to assess the role of SMFD-score predicting major chronic diseases independently from age, sex, fat area at 66% of tibia length, and multimorbidity (Table 4). Higher SFMD-scores were inversely associated with prevalence and incidence of: Parkinson’s disease (OR = 0.91; 95% CI: 0.85-0.96; p = 0.002), Stroke (OR = 0.88; 95% CI: 0.83-0.93; p < 0.001), Hip osteoarthritis (OR = 0.95; 95% CI: 0.91-0.99; p = 0.02). No significant associations were observed for myocardial infarction, angina, heart failure, or cancer.

**Table 4:** GEE, ordinal repeated measures, with a multinomial distribution. Incident and prevalent diseases compared to those subjects that do not report the specific disease, predicted by SFMDscore. The models consider also age, sex, fat area at 66% of tibia length, and multimorbidity.

|  | OR | 95%CI | p-value |
| --- | --- | --- | --- |
| Metabolic syndrome | 1.03 | 0.98-1.07 | 0.18 |
| **Hip arthritis** | 0.95 | 0.91-0.99 | 0.02 |
| Knee arthritis | 0.98 | 0.95-1.02 | 0.31 |
| Peripheral arterial disease | 0.99 | 0.95-1.03 | 0.63 |
| **Parkinson's disease** | 0.91 | 0.85-0.96 | 0.002 |
| **Stroke** | 0.88 | 0.83-0.93 | <0.001 |
| Congestive heart failure | 0.99 | 0.96-1.04 | 0.83 |
| Myocardial infarction | 1.05 | 0.98-1.11 | 0.08 |
| Angina pectoris | 1.02 | 0.96-1.08 | 0.43 |
| Chronic liver disease | 1.03 | 0.96-1.11 | 0.40 |
| Hypertension | 0.99 | 0.96-1.04 | 0.99 |
| Cancer | 1.02 | 0.96-1.09 | 0.46 |

## 4.0 Discussion

The main results of this longitudinal analysis of the InCHIANTI cohort demonstrated that the SFMD-score proved to be a comprehensive and clinically meaningful tool to characterize the progressive deterioration of muscle function in older adults. The decline in SFMD was paired by an increasing burden of disability (both in BADL as in IADL), multimorbidity score, frailty syndrome, hospitalizations, number of falls, and chronic diseases, reinforcing the tight association between muscular resilience and global health status in aging. According to theoretical conceptualization and clinical manifestation on sarcopenia, our findings underscore that muscle dysfunction in older adults extends far beyond simple reductions in muscle mass.

Although reductions in muscle area were evident, the concomitant losses in muscle strength, lower leg muscle power, and muscle density point to qualitative deteriorations in muscle tissue that probably could precede and outpace purely anatomical changes (Freitas et al., 2024). This dissociation between mass and function is now well-recognized in the conceptual evolution of aging muscle components (Chang et al., 2017). In particular, the rapid decline in lower leg muscle power as captured by the Sankey flow diagrams, that analyses during the study the changes in quintile of distribution in the quality and quantity of the muscle, supports the evidence that reductions in contraction velocity and neuromuscular efficiency represent early and highly sensitive indicators of functional impairment (Radaelli et al., 2023). As matters of fact, SMFD could better predict functional dependency than either mass or strength alone, likely reflecting its reliance on both central (neurological) and peripheral (muscular) mechanisms (Pellegrino et al., 2025). The SFMD-score as proposed integrates these functional and morphological dimensions of the muscle, thereby capturing the complex biological heterogeneity that underlies age-related muscle dysfunction.

Unlike traditional definitions focused primarily on lean mass and/or strength, the SMFD theoretical framework incorporates potential contributors such as myosteatosis (Miljkovic and Zmuda, 2010), inflammation (Kahn et al., 2022; Paganelli and Di Iorio, 2025), neural activation deficits (Pellegrino et al., 2024), and changes in muscle composition and architecture (Santanasto et al., 2023). Our results demonstrate that higher SFMD-scores provide protection from multiple clinically relevant and adverse endpoints including basic and instrumental activities of daily living disability, poor physical performance, frailty phenotype, hospital admissions, and falls even adjusting for age, sex, multimorbidity, and adiposity.

Notably, the association between higher SFMD and reduced incidence/prevalence of chronic diseases, particularly Parkinson’s disease, stroke, and hip osteoarthritis, suggests that muscular dysfunction may precede or reflect early subclinical stages of systemic disease processes. While causality cannot be inferred categorically, these findings suggest the possibility that interventions aimed at preventing muscle decline may have broader protective role beyond merely the musculoskeletal aging trajectories effects. From a clinical perspective, these results reinforce the potential utility of incorporating SMFD-based assessments into routine geriatric evaluations. The ability of SFMD to stratify risk across multiple domains makes it a promising candidate for guiding early preventive strategies, tailored exercise prescriptions, and more targeted rehabilitation interventions.

Moreover, the observed resilience among a small subset of participants who maintained high SFMD scores throughout follow-up warrants further exploration. Understanding the biological, behavioral, and possibly genetic determinants of this “high-functioning aging” phenotype may yield insights into mechanisms of muscle preservation and healthy longevity.

Several strengths of this study merit emphasis, including its longitudinal design, large sample size, comprehensive phenotyping, and integration of multiple muscle-related domains into a unified functional score. However, limitations must also be acknowledged. The main limitation of this study concerns the devices used to calculate the SMFD score, which are rarely available for routine clinical assessment and are generally limited to specialized research facilities. Moreover, the evaluations performed are costly in terms of both time and financial resources. Nonetheless, the operational implications of an integrated assessment of the various muscle components may support an early and personalized clinical approach to the consequences of muscle aging.

Moreover, our observational design precludes definitive causal inference; survivorship bias may have led to underestimation of associations; and the SFMD algorithm, while comprehensive, may require external validation in independent populations and diverse ethnic groups.

In conclusion, our findings highlight the clinical value of adopting the broader framework of Skeletal Muscle Function Deficit (SMFD) to better capture the multidimensional nature of muscle dysfunction in aging. The SFMD score emerges as a powerful and flexible tool for predicting a wide range of adverse outcomes in older adults, with potential applications in clinical care, prevention, and research.

## Data Availability

All data produced in the present study are available upon reasonable request to the authors

